# Algorithmic Versus Expert Rankings of Large Language Models in Peritoneal Dialysis Prescription Review: A Trap-Embedded Synthetic Benchmark

**DOI:** 10.64898/2026.05.28.26354383

**Authors:** Chao-Hsuan Wei, Hsuan-Jen Lin, Wu-Wei Lai, Hsuan-Ming Lin

## Abstract

**Background:** Clinical LLM benchmarks rarely test whether algorithmic rankings agree with expert clinical judgment. We developed a trap-embedded peritoneal dialysis (PD) benchmark comparing multiple scoring constructs with blinded nephrologist ratings.

**Methods:** We generated 125 synthetic PD cases containing 13 ISPD-aligned trap types. Five LLMs (Claude Sonnet 4.5, GPT-5.4, Gemini 3.1 Pro, DeepSeek-R1, Grok 4.1 Fast) evaluated each case three times at temperature 0 (1,875 calls). Primary outcome was must-identify TDR_must_, analyzed with GEE and case-clustered bootstrap. Secondary analyses included a verbosity-sensitive alarm-burden proxy, WCS, relaxed-match scoring, WCS sensitivity analyses, and a 25-output blinded expert adequacy substudy. Must-identify kappa was 0.89 in Stage 1 and 0.92 in Stage 2.

**Results:** Rankings were discordant. Recall ranked Claude (0.977) and GPT-5.4 (0.955) above the other models (0.86–0.90, p<0.0001). The alarm-burden proxy favored concise models (Grok 0.689; 21.6 vs 2.4 issues/case), while WCS produced a third ordering. In the expert substudy, inter-rater concordance was strong (rho 0.977), but WCS did not show a positive association with expert adequacy (rho -0.17, p=0.41).

**Conclusion:** Clinical LLM rankings in PD prescription review depend strongly on scoring construct. Algorithmic metrics should be reported alongside blinded expert adequacy ratings and should not alone determine deployment.

## 1 Introduction

Peritoneal dialysis (PD) prescription review is a multi-domain clinical task increasingly proposed for large language model (LLM) augmentation, yet rigorous benchmarks of LLM performance in this setting are lacking. PD prescriptions are continuously adjusted in response to peritoneal equilibration test results, residual kidney function trajectory, ultra-filtration profile, adequacy targets, and patient tolerance [1–4]. Persistent clinical pitfalls — failure to adapt long dwells in high transporters, missed reassessment after peritonitis, continued underdialysis without modality revision, and reliance on solute targets without clinical contextualization [5–8] — make PD prescription review a demanding multi-step adequacy task that an algorithmic decision aid may be tempted to summarize with a single performance number.

LLMs are increasingly evaluated in medical settings, but many studies rely on single algorithmic metrics or knowledge-style benchmarks [9, 10]. For PD prescription review, this raises a construct-validity concern: a metric may be reproducible yet fail to measure clinically meaningful adequacy [11]. PD prescription review is a useful stress test because safe assessment requires reasoning across solute clearance, volume control, transporter status, residual kidney function, prescription burden, and treatment strategy.

The nephrology LLM literature remains weighted toward education, patient-facing explanations, and general knowledge assessment [9, 12]. AI and machine-learning studies in PD have more often focused on prediction, prognosis, or digital-twin concepts than on clinician-facing review of chronic PD prescription adequacy [13–15]. A useful benchmark for PD prescription review must test both trap recognition and the burden of low-value issue generation.

Five contemporary LLMs were therefore benchmarked on a trap-embedded synthetic PD case set. The primary objective was to compare model rankings across recall, an exploratory alarm-burden proxy (verbosity-sensitive, not adjudicated precision or F1), WCS, and expert global adequacy ratings. The secondary objective was to assess whether an algorithmic WCS aligned with blinded nephrologist judgment. The expert substudy was framed as an anchoring exercise rather than a definitive validation study because the sample was intentionally modest and designed to detect major construct-validity discordance.

## 2 Methods

### 2.1 Study Design and Ethics

This was a cross-sectional benchmark study using fully synthetic PD prescription cases (overall study design in Figure 1). No real patient records, identifiable private information, biospecimens, or patient-clinician interactions were used. Clinician authors independently reviewed synthetic cases and model outputs as part of benchmark construction and adjudication. The broader research program of which this synthetic benchmark forms the first phase is approved under institutional review board protocol 113TMANH-REC011(CR-1) (Tainan Municipal An-Nan Hospital Institutional Review Board); the present synthetic phase falls within the approved scope.

**Figure 1.**
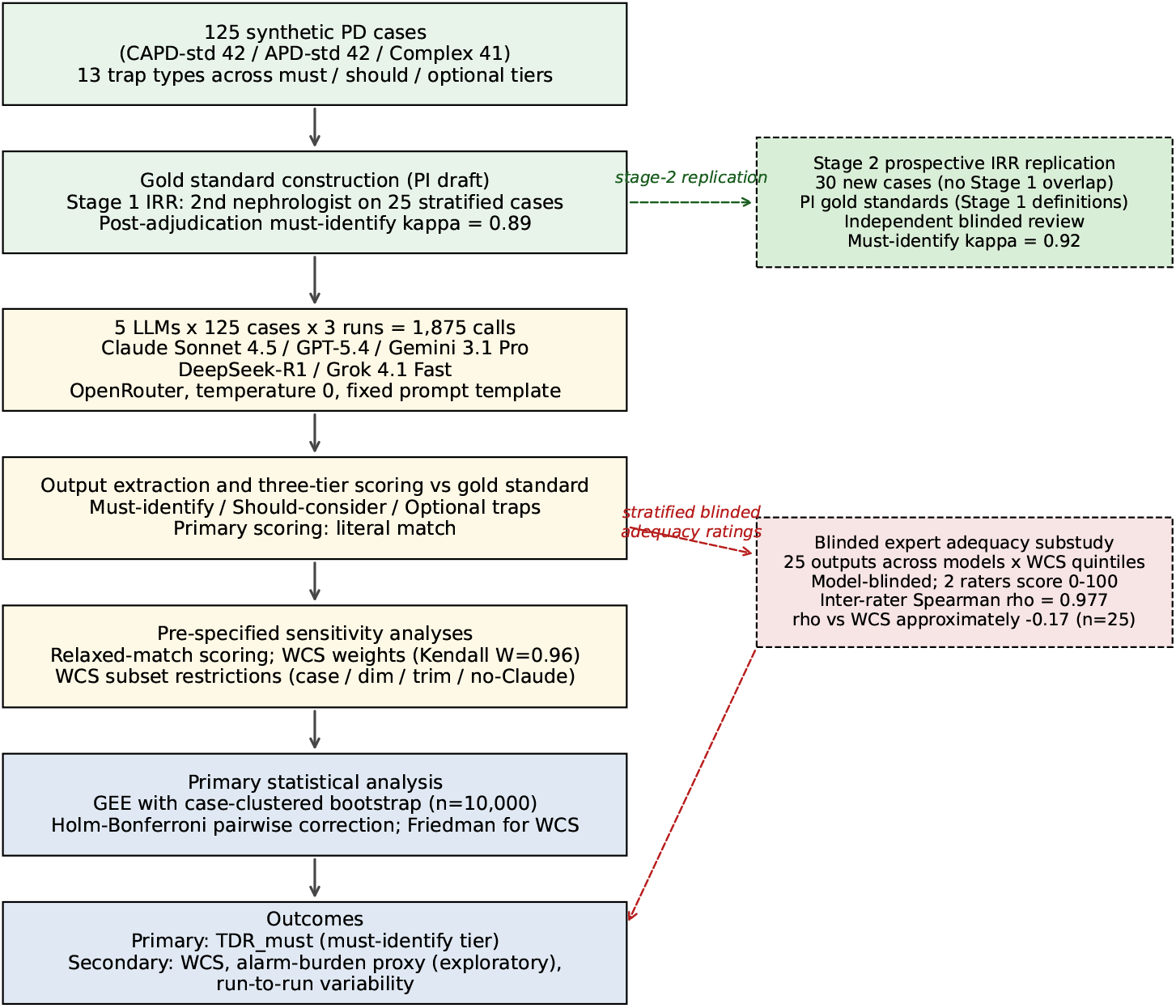
Study pipeline. Main flow (vertical, 7 boxes): synthetic case generation; gold-standard construction with Stage 1 inter-rater reliability (must-identify kappa 0.89); LLM evaluation (1,875 calls); output extraction and three-tier scoring; pre-specified sensitivity analyses; primary statistical analysis; and outcomes. Side branches (dashed, right): Stage 2 prospective IRR replication on 30 non-overlapping cases (must-identify kappa 0.92); and the blinded expert adequacy substudy (25 stratified outputs, two nephrologists, 0–100 scale; inter-rater rho 0.977, rho versus WCS approximately -0.17).

The analysis was designed as the synthetic phase of a broader PD LLM evaluation program. This manuscript reports only the synthetic benchmark. No claims are made about performance on real patient records or about autonomous clinical use.

### 2.2 Case Generation

We generated 125 synthetic PD cases stratified into CAPD-standard, APD-standard, and complex modality groups. Cases were assembled programmatically from a parameterised template covering demographics, comorbidities, prescription parameters, PET results, longitudinal laboratory data, medications, and residual kidney function trajectories; clinician authors reviewed and edited each case for clinical plausibility before lock. Thirteen trap types were embedded across cases, spanning solute clearance, transporter-dwell mismatch, ultrafiltration failure, residual function decline, modality strategy, volume overload, electrolyte management, prescription burden, and biocompatibility. The trap-embedded design was informed by adversarial and rubric-based medical LLM evaluation frameworks [16–19].

Cases were clinically plausible rather than patient-derived; the synthetic design embedded predefined PD safety pitfalls without exposing patient data. Embedded traps were organized into must-identify, should-consider, and optional tiers; inferential claims were restricted to the must-identify tier because it achieved the strongest inter-rater reliability. Trap definitions and gold standards were finalized before model scoring, and the LLM prompts did not disclose the trap types or counts embedded in each case. Complex cases included recent peritonitis, modality switch, or post-transplant return scenarios. Case characteristics are summarized in Table 1; detailed trap definitions are in Supplementary Appendix S1.

**Table 1.** Case characteristics by modality group. Values are median (IQR) unless otherwise noted.

| Characteristic | CAPD | APD | Complex | Overall |
| --- | --- | --- | --- | --- |
| n | 42 | 42 | 41 | 125 |
| Age, years | 51.0 (40.0–58.8) | 52.5 (44.0–62.8) | 49.0 (47.0–61.0) | 51.0 (44.0–61.0) |
| Female, n (%) | 16 (38) | 13 (31) | 19 (46) | 48 (38) |
| BMI, kg/m<sup>2</sup> | 22.4 (20.4–24.6) | 23.5 (22.2–26.1) | 24.0 (21.1–26.0) | 23.3 (21.1–25.9) |
| PD vintage, months | 18.0 (8.0–27.0) | 14.0 (7.0–20.8) | 13.0 (5.0–33.0) | 15.0 (7.0–24.0) |
| Anuric, n (%) | 3 (7) | 4 (10) | 4 (10) | 11 (9) |
| Residual eGFR, mL/min | 2.8 (1.3–3.7) | 2.6 (1.1–4.0) | 2.8 (1.1–4.0) | 2.7 (1.2–3.8) |
| Urine output, mL/day | 427 (265–625) | 403 (293–688) | 370 (215–765) | 400 (241–722) |
| Total weekly Kt/V | 1.9 (1.8–2.1) | 1.9 (1.7–2.1) | 1.9 (1.7–2.1) | 1.9 (1.7–2.1) |
| D/P creatinine 4h | 0.70 (0.60–0.80) | 0.70 (0.50–0.80) | 0.70 (0.60–0.80) | 0.70 (0.50–0.80) |
| Icodextrin used, n (%) | 3 (7) | 16 (38) | 9 (22) | 28 (22) |
| Embedded traps per case | 2 (2–3) | 3 (2–3) | 2 (2–3) | 2 (2–3) |

### 2.3 Gold Standard Construction and Reliability

The PI drafted the initial gold standards from the predefined trap definitions. Each gold standard specified embedded traps, severity tier, expected detection text, expected action, guideline basis, key data points, and a four-dimension scoring rubric. A second nephrologist independently reviewed a stratified random sample of 25 cases. After adjudication and operational refinement of ambiguous trap definitions, aggregate trap-identification Cohen’s kappa was 0.76 and must-identify tier kappa was 0.89. To address potential overfitting during this Stage 1 adjudication, a Stage 2 prospective reliability replication was conducted on 30 newly generated non-overlapping cases using the finalized operational definitions (results in §3.2).

### 2.4 LLM Evaluation

Five contemporary LLMs were evaluated through a unified API using a fixed case prompt and system prompt. Each case-model pair was queried three times at temperature 0, yielding 1,875 total calls. The prompt requested assessment, issues, recommendations, and reasoning sections. Outputs were parsed before scoring, with incomplete sections scored from available text according to the pre-specified extraction rules.

The evaluated models were Claude Sonnet 4.5, GPT-5.4, Gemini 3.1 Pro, DeepSeek-R1, and Grok 4.1 Fast. All calls used the same prompt template, temperature, and token limit. Model names refer to API identifiers and returned snapshot metadata recorded at inference time; the routing configuration, fallback policy, and two retried calls (out of 1,875) are reported in Supplementary Table S2. The model snapshot should be interpreted as time-specific.

### 2.5 Scoring and Statistical Analysis

The primary endpoint was case-level TDR_must_, defined as the proportion of must-identify traps detected in each case. The unit of analysis was the case-model-run observation among cases containing at least one must-identify trap (93 of 125 cases; 1,395 observations across 5 models × 3 runs). Generalized estimating equations with robust standard errors were used for model comparisons, and pairwise contrasts were estimated with case-clustered nonparametric bootstrap (n=10,000 resamples; Monte Carlo resolution 0.0001) with Holm-Bonferroni correction; resampling was performed at the case level and preserved all model and run observations within each resampled case. P-values at the bootstrap floor are reported as *<* 0.001.

Secondary analyses included a verbosity-sensitive alarm-burden proxy (trap yield per LLM-raised issue), WCS, scoring-strictness sensitivity, run-to-run variability, and the blinded expert adequacy rating substudy. The proxy used the number of LLM-raised issue items as the denominator (rather than manually adjudicated false positives) and is therefore reported as exploratory rather than as adjudicated precision or F1.

WCS combined safety, diagnostic, treatment, and reasoning dimensions. For the blinded expert adequacy rating substudy, 25 run-1 outputs were stratified by (model × WCS quintile; seeded selection, seed 2026) and rated 0–100 by two board-certified nephrologists blinded to model identity, gold standard, and all algorithmic scores. One rater (HML) had constructed the gold standards and the other (HJL) had served as the independent Stage 1/Stage 2 IRR adjudicator; this prior involvement is considered in interpretation. Spearman correlation quantified inter-rater agreement and the association between expert ratings and WCS. Triplicate runs were retained because deterministic LLM settings can still show output variability [20, 21].

WCS was a weighted composite of four dimensions (Safety, Diagnostic accuracy, Treatment appropriateness, Reasoning quality). Each dimension was scored on a 0–5 raw-point scale using per-case rubric criteria (positive and negative), clipped at [0, 5], normalized by the dimension cap of 5, and aggregated:

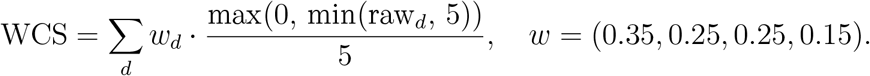

WCS is bounded in [0, 1]; raw per-dimension scores in dimension-breakdown analyses retain the 0–5 scale. The full rubric is in Supplementary Appendix S5. Scoring-strictness sensitivity re-ran trap detection under a relaxed lexical-match threshold.

## 3 Results

### 3.1 Case Characteristics

The 125 synthetic cases were balanced across modality groups and contained a median of 2 embedded traps per case (Table 1).

All 1,875 model calls completed. First-attempt success was 99.89%, and two transient failures were retried successfully. Parse success was 99.8%; four outputs were truncated mid-section but contained sufficient content for partial scoring under the extraction rules. Extraction validation passed for 98.5% of outputs with all four requested sections complete.

### 3.2 Gold Standard Reliability and Stage 2 Prospective Replication

After adjudication on the Stage 1 sample of 25 cases, aggregate trap-identification Cohen’s kappa was 0.76 (95% CI 0.71–0.81) over 325 trap-case cells, with 88.6% raw agreement. Reliability differed substantially by tier: must-identify kappa 0.89 (96.0% agreement), should-consider kappa 0.27, and optional kappa 0.19. Inferential analyses were therefore restricted to must-identify traps, with lower tiers treated descriptively.

Because Stage 1 adjudication used the same cases for both definition refinement and validation, a Stage 2 prospective replication was conducted on 30 newly generated cases (seed=2027; no overlap with Stage 1; identical modality stratification). The PI built gold standards on the new cases using the Stage 1 operational definitions, and the second nephrologist independently reviewed all 30 cases blinded to the gold standard. Aggregate trap kappa was 0.77 with must-identify kappa 0.92, equal to or exceeding the Stage 1 must-identify value. Should-consider and optional tiers remained limited (kappa 0.67 and 0.14, respectively, with persistent R2 under-coding of T13), so inferential conclusions remain restricted to must-identify traps. Per-trap reliabilities and per-case agreement are in Supplementary Appendix S1.

### 3.3 Primary Recall Ranking

Models differed significantly on TDR_must_ among the 93 cases containing at least one must-identify trap. Marginal means were 0.977 for Claude, 0.955 for GPT-5.4, 0.899 for DeepSeek, 0.864 for Gemini, and 0.864 for Grok. The omnibus Wald chi-square was 53.0 (p<0.0001). After Holm-Bonferroni correction, Claude and GPT-5.4 each significantly outperformed Gemini, DeepSeek, and Grok. Claude versus GPT-5.4 was not significant after multiplicity correction, and the lower three models did not differ significantly from each other.

Per-trap separation was most visible for residual kidney function decline (T07), a trap that required interpreting both a numerical decline rate and clinical significance. The largest model spread, however, occurred on persistent hyperkalemia without systematic prescription evaluation (T08), where detection rates ranged from Claude 0.88 and GPT-5.4 0.74 down to Gemini 0.49, DeepSeek 0.50, and Grok 0.32 (Figure 3). T08 explicitly required the model to recognize when systematic evaluation of diet, constipation, acidosis, residual kidney function, and PD prescription parameters had been omitted before reaching for potassium binders or HD conversion; concise models tended to acknowledge hyperkalemia and recommend treatment without flagging the missing systematic workup. By contrast, several high-salience must-identify traps approached ceiling performance in the highest-recall models, limiting their ability to discriminate among top performers.

### 3.4 An Alarm-Burden Proxy Inverts the Ranking (Exploratory)

We also computed a verbosity-sensitive alarm-burden proxy using the number of LLM-raised issue items as the denominator. Because this denominator includes both true false positives and legitimate clinical observations outside the embedded trap set, we report the metric (trap yield per raised issue) rather than as adjudicated precision or F1, and treat the resulting ranks as exploratory signals. Under this metric the ranking inverted (Friedman omnibus p<0.001). Mean issues raised per case were 21.6 (Claude), 13.0 (GPT-5.4), 10.1 (Gemini), 4.1 (DeepSeek), and 2.4 (Grok); corresponding proxy values were 0.142, 0.201, 0.232, 0.567, and 0.689. The two highest-recall models had the lowest proxy values and highest issue counts; the two lowest-recall models achieved the highest proxy values with concise outputs. The rank inversion mirrors the 21.6-versus-2.4 issues-per-case gap, a pattern consistent with verbosity rather than detection accuracy. Without false-positive adjudication, these rank differences signal alarm burden, not fewer clinical errors.

### 3.5 Weighted Clinical Score Yields a Third Intermediate Ordering and Remains Stable under Sensitivity Analysis

WCS produced an intermediate ranking that differed from both recall and the alarm-burden proxy. Mean WCS was 0.665 for Claude, 0.581 for GPT-5.4, 0.498 for Gemini, 0.486 for DeepSeek, and 0.448 for Grok (Friedman p<0.001; mean ranks 1.45, 2.45, 3.50, 3.52, 4.08). Under relaxed lexical matching, TDR_must_ values converged toward ceiling (0.983–1.000) and the omnibus difference disappeared (p=0.27), whereas WCS remained discriminative with the same broad ordering (Figure 2). Run-to-run disagreement was observed despite temperature 0: at least one inter-run TDR_must_ disagreement occurred in 8.6% of Claude, 16.1% of GPT-5.4, 26.9% of Gemini, 29.0% of DeepSeek, and 30.1% of Grok case-model pairs, so triplicate sampling was necessary to estimate benchmark behavior under routine serving-stack variability. WCS dimension breakdown showed the largest model spread on Reasoning (Claude 2.91 vs DeepSeek 1.05; ratio 2.8×) despite this dimension carrying the lowest weight (0.15); the most compressed range was on Diagnostic accuracy (3.71 vs 2.83; 1.3×).

**Figure 2.**
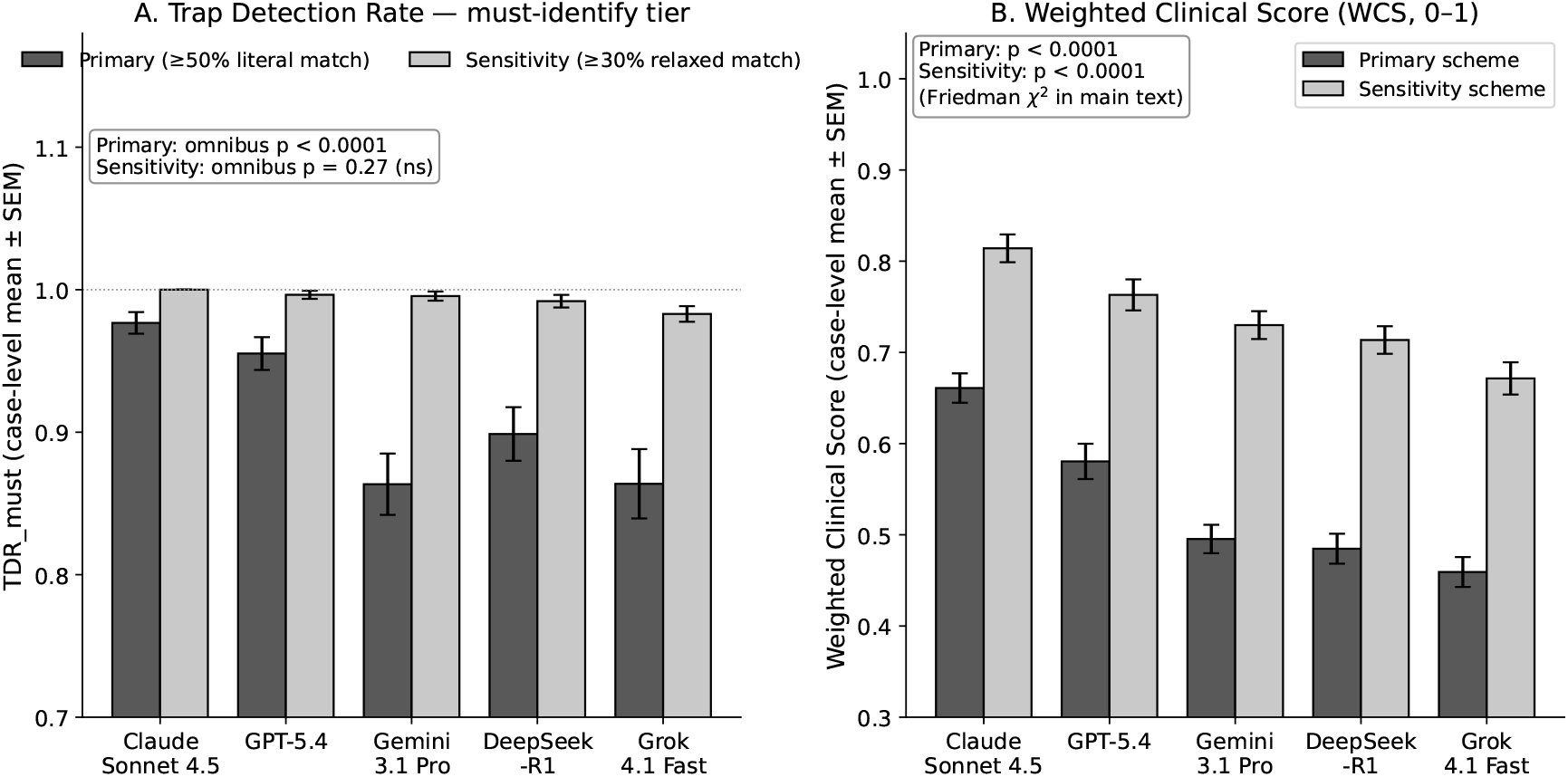
Per-model TDR_must_ (recall) under primary literal-match scoring and pre-specified relaxed-match sensitivity scoring. Bars show marginal mean TDR_must_ (93 must-identify-containing cases ×5 models ×3 runs). Under primary scoring the omnibus difference is significant (*p <* 0.001); under relaxed scoring it disappears as values converge toward the ceiling (*p* = 0.27).

**Figure 3.**
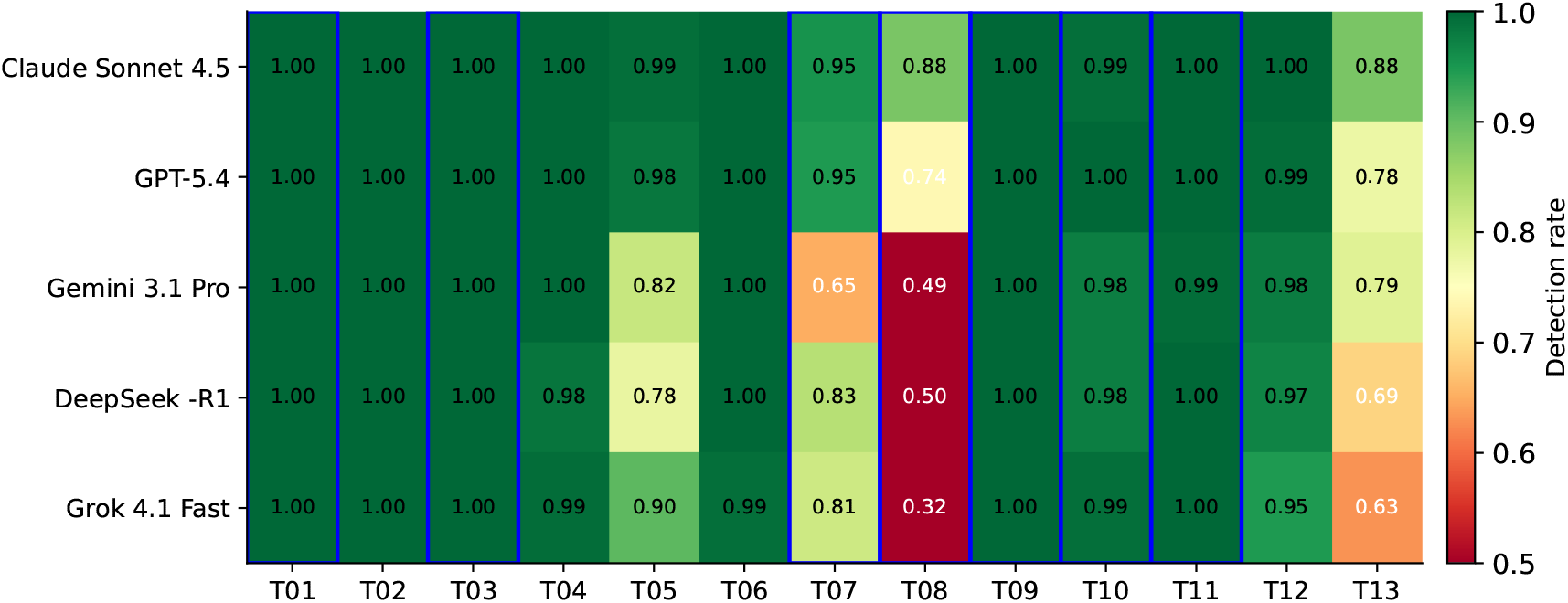
Per-trap detection rate by model under primary literal-match scoring. Must-identify-tier traps (T01, T03, T07–T11) are outlined in blue; the remaining columns are should-consider (T02, T04, T05, T12) or optional (T06, T13) tiers retained descriptively. Color scale: red (low) to green (high). The largest cross-model spread occurs on T08 (persistent hyperkalemia without systematic evaluation), from 0.32 (Grok) to 0.88 (Claude).

A weighting-scheme sensitivity analysis confirmed that the WCS ranking does not depend on the primary 0.35/0.25/0.25/0.15 weight choice (Supplementary Appendix S6). Across four pre-specified schemes (primary; equal 0.25 each; safety-heavy 0.50/0.20/0.20/0.10; reasoning-up 0.30/0.20/0.20/0.30), Kendall’s W=0.963 (p=0.004); Claude was rank 1, GPT-5.4 rank 2, and Grok rank 5 under every scheme, with only a single adjacent swap between DeepSeek and Gemini under safety-heavy weighting. Spearman correlation between WCS and the blinded expert mean rating remained near zero under every scheme (-0.083 to -0.173; all 95% CIs included zero), so the WCS-versus-expert dissociation is not a weighting artifact.

### 3.6 Blinded Expert Adequacy Rating Substudy

Two nephrologists independently rated 25 blinded outputs sampled across models and WCS quintiles. Their ratings were strongly concordant (Spearman rho 0.977, p<0.001). However, WCS did not correlate with either rater’s score or with the mean expert rating (mean-rating rho -0.17, p=0.41; Figure 4). Several low-WCS outputs received high expert ratings, and several high-WCS outputs received only moderate expert ratings. Given n=25 and ratings compressed at the upper end of the scale, this should be read as failing to demonstrate a positive WCS–expert association rather than as definitive null evidence (see Limitations). Pre-specified subset sensitivity analyses (restricting to must-identify-trap cases, to the Safety dimension only, to trimmed expert ratings, or excluding Claude outputs) yielded Spearman *ρ* values from −0.32 to +0.01, with all 95% CIs including zero; no subset analysis showed a positive WCS–expert association (Supplementary Appendix S7).

**Figure 4.**
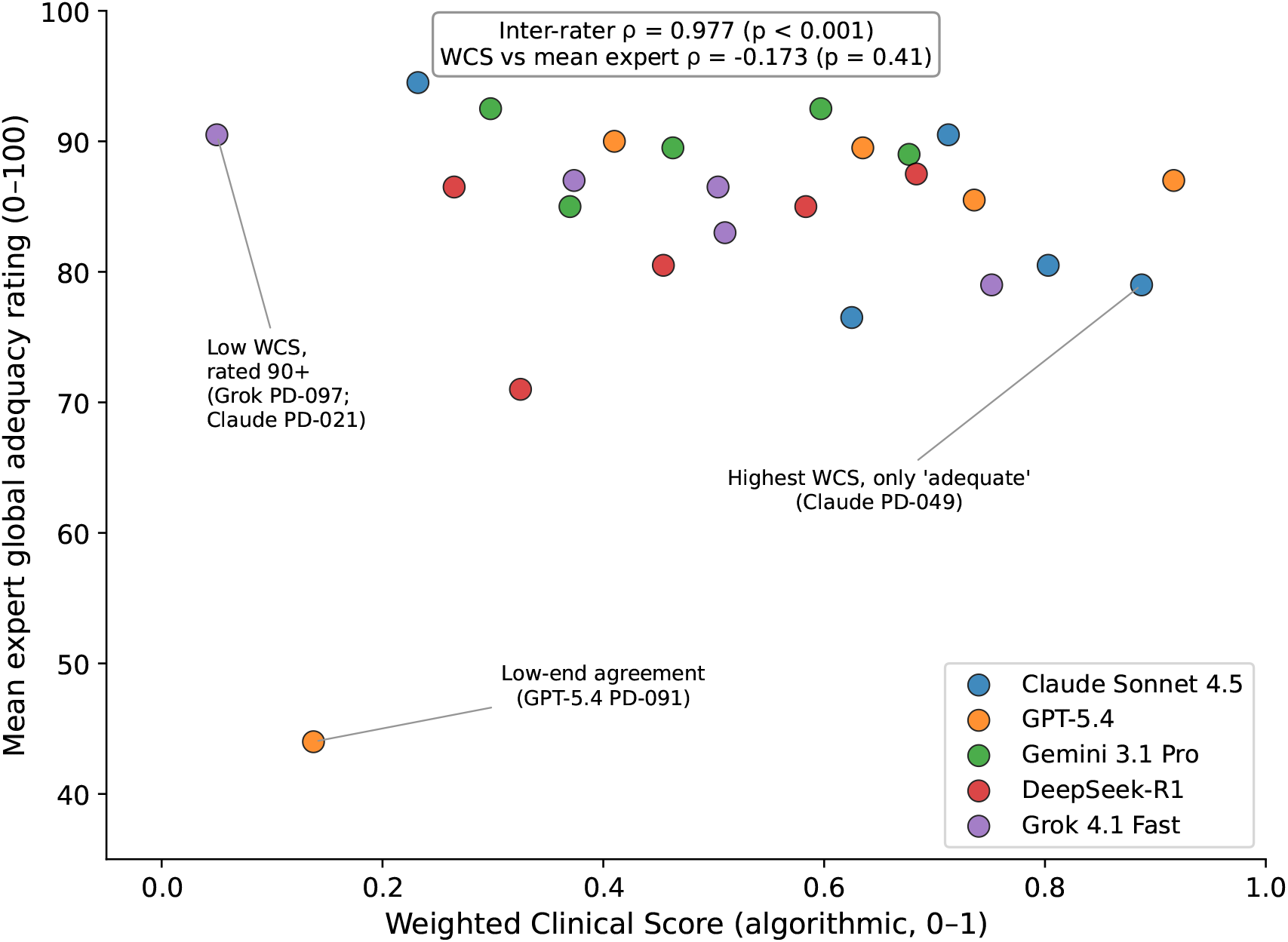
Association between WCS and blinded expert global adequacy ratings in the stratified expert-rated substudy (n=25 outputs sampled across models and WCS quintiles; Spearman rho approximately −0.17). Selected mismatch cases are annotated.

## 4 Discussion

### 4.1 Principal Findings: Algorithmic Rankings Differ from Each Other and from Expert Judgment

Three algorithmic rankings of the same five LLMs were discordant on the same 1,875-call dataset. Recall favored Claude and GPT-5.4 well above the other three. The exploratory alarm-burden proxy reversed this, favoring Grok and DeepSeek because verbose models raised many more issues per case (21.6 versus 2.4); we read this as a verbosity signal rather than as adjudicated precision. WCS yielded an intermediate ordering favoring Claude and GPT-5.4. A small stratified blinded expert-anchoring substudy further suggested that WCS may not capture global clinical adequacy: the two nephrologists agreed near-perfectly with each other (Spearman rho 0.977) but the analysis did not demonstrate a positive association between either rater’s score and WCS (combined rho approximately −0.17, p=0.41).

This dissociation is the key construct-validity probe. Two scoring directions account for it: WCS rewarded literal multi-criterion lexical coverage, while expert ratings weighed whether the principal trap was correctly identified and prioritized. WCS is therefore highly reliable as a computation but did not establish construct validity in this small stratified validation sample (n=25, two raters). Representative mechanistic case audits illustrating both directions are provided in Supplementary Box S1.

### 4.2 Clinical Implications: Algorithmic Rankings Are Not Substitutes for Expert Judgment

The algorithmic rankings remain valid descriptions of metric-specific behavior: Claude emits more keyword-overlap matches than Grok, Grok raises fewer issues per case, and WCS scores Claude higher than Grok. However, none of these rankings predicted blinded expert adequacy ratings in this validation sample. A clinician relying on a single algorithmic metric could reach a model-selection conclusion not supported by blinded expert adequacy ratings in this substudy. Three implications follow: (a) algorithmic LLM benchmarks in clinical domains should be reported alongside expert validation rather than as substitutes [11]; (b) high recall is not equivalent to clinical thoroughness, since flagging 13–22 issues per case generates alarm fatigue without commensurate gold-standard detection and was not rated higher by experts than concise models that flagged 2–4 issues; and (c) reliability does not imply validity, even within a multi-dimensional rubric.

For must-identify traps, the gold standard was highly reliable across Stage 1 and Stage 2 (kappa 0.89 and 0.92), so recall rankings and per-trap separation on T07 and T08 are inferentially supported. For should-consider and optional traps, inter-rater reliability was insufficient (especially persistent T13 under-coding), so model comparisons within those tiers are descriptive only. The WCS-versus-expert dissociation is the most robust qualitative finding because it does not depend on lower-tier trap reliability, although its magnitude (rho approximately −0.17) is constrained by the small expert sub-study.

### 4.3 Methodological Strengths

The trap-embedded design anchors evaluation to specific ISPD-guideline-aligned pitfalls rather than to open-ended rubrics, and generator–evaluator separation reduces self-preference bias. Stage 2 prospective replication on 30 newly generated cases (no overlap with Stage 1) confirmed must-identify kappa of 0.92, indicating that the refined trap definitions are not artifacts of post-hoc calibration on the Stage 1 sample. Pre-specifying recall, an exploratory alarm-burden proxy, WCS, and a scoring-strictness sensitivity analysis on the same dataset exposes metric-dependence that a single-metric benchmark would have hidden. Blinded expert anchoring with inter-rater Spearman rho probes construct validity, addressing the absence of expert-rated adequacy checks in many medical-LLM benchmarks. Reporting follows principles aligned with the TRIPOD-LLM guideline for studies using large language models [22], with adaptations for this synthetic benchmark design noted in the upload-package reporting note.

### 4.4 Limitations

The benchmark used synthetic English-only cases; the evaluated model versions represent a time-stamped snapshot, and the full pipeline is archived for reproducibility against future model releases. The alarm-burden proxy denominator counts all LLM-raised issues, including clinically legitimate observations outside the embedded trap set; the resulting ranking inversion describes alarm density per case rather than adjudicated false-positive rates, and does not imply that concise models commit fewer clinical errors. The primary recall ranking may also partly reflect scoring-threshold sensitivity given the ceiling effect under relaxed matching (Section 3.5).

The expert adequacy substudy used only 25 stratified outputs with ratings compressed at the upper end of the 0–100 scale, and both raters were investigator-raters performing blinded output assessment; the point estimate of rho approximately −0.17 is consistent with the absence of a positive WCS–expert association in this sample but cannot exclude all positive association, and replication with independent external raters and a larger panel is needed. Inter-rater reliability for the should-consider and optional tiers was insufficient (especially T13 under-coding), so inferential conclusions are restricted to the must-identify tier. Hallucination analyses remain exploratory pending final adjudication of trigger boundaries [23, 24], and run-to-run variability at temperature 0 is characterized in the supplementary methods [20, 21].

The synthetic design also omits aspects of real-world PD review that future work must address. Two are most clinically consequential: longitudinal trends across repeated PET, residual kidney function, and Kt/V measurements over months to years (for example, drift in ultrafiltration adequacy that triggers incremental dosing transitions); and patient-side and multi-disciplinary inputs (for example, modality switch driven by exchange burden, family caregiving capacity, or input from the renal dietitian and home-care coordinator). The English-only corpus also cannot capture symptom-reporting and adherence narratives in other languages. Results may further depend on the specific prompt template, output-format instructions, and API serving stack used in this benchmark; alternative prompts could change the issue-density and dimension scores reported here. The trap-embedded synthetic design is best regarded as a controlled methodological foundation; deidentified real-world cohort validation (planned in our subsequent real-world validation study) is required before any clinical deployment recommendation.

## 5 Conclusion

This trap-embedded synthetic benchmark shows that model rankings are metric-dependent: recall, an alarm-burden proxy, and WCS yielded discordant orderings on the same outputs. WCS was internally stable across weighting schemes (Kendall’s W=0.96) but in a small stratified blinded expert-anchoring substudy (n=25) did not demonstrate a positive association with expert adequacy ratings (combined rho approximately −0.17); confirmation in a larger expert panel is needed. Clinical LLM benchmarks should therefore report multi-axis algorithmic metrics together with an expert-anchored clinical adequacy assessment, rather than rely on any single algorithmic ranking for deployment decisions. No model evaluated here should be selected for PD prescription-review deployment based on these algorithmic rankings alone.

## Data Availability

All data produced in the present study are available upon reasonable request to the authors

## Author Contributions

CHW: Writing – original draft, clinical interpretation, manuscript revision, first author. HJL: Independent gold standard review, writing – review and editing. WWL: Writing – review and editing. HML: Study conception, design, methodology, statistical design, gold standard construction, supervision, writing – review and editing, corresponding author.

## Use of AI and AI-Assisted Technologies

The authors used AI-assisted tools (large language model assistants, including ChatGPT and Claude) for author-supervised language editing, translation between Traditional Chinese and English, drafting assistance, LaTeX formatting, and internal consistency review of the manuscript. All study design decisions, statistical analyses, interpretation of results, scientific claims, and final wording were verified and approved by the authors, who take full responsibility for the content of the manuscript. The five large language models evaluated in this study (Claude Sonnet 4.5, GPT-5.4, Gemini 3.1 Pro, DeepSeek-R1, Grok 4.1 Fast) were used solely as the experimental object of the benchmark and did not contribute to writing.

## Ethics Approval

This study used only fully synthetic cases and did not involve real patient records, identifiable private information, biospecimens, or patient-clinician interactions. The broader research program is approved under institutional review board protocol 113TMANH-REC011(CR-1) (Tainan Municipal An-Nan Hospital Institutional Review Board); the present synthetic phase falls within the approved scope.

## Data Availability

Materials supporting this benchmark are organized in three tiers. (i) **Synthetic cases, prompts, scoring rubrics, and reproducibility code** will be made available in the project repository upon publication; a private reviewer-access link will be provided on request during peer review. (ii) **Raw API responses** (request payloads, response bodies, returned model identifiers, token usage, and finish reasons) are archived locally with model identifiers and run metadata. Because raw provider outputs are subject to OpenRouter and underlying provider terms of service (Anthropic, OpenAI, Google, xAI, DeepSeek), they will be released to non-commercial academic researchers upon request after publication, under a data use agreement (DUA) that limits redistribution and prohibits use of these outputs to train competing AI systems. (iii) **Real-world patient data** are not part of this study and are not released from this repository. A run manifest records model identifiers, returned snapshot IDs, API parameters, retry policy, and release status.

