## Supplementary for "Algorithmic Versus Expert Rankings of Large Language Models in Peritoneal Dialysis Prescription Review: A Trap-Embedded Synthetic Benchmark"

### Supplementary Material: Metric-Dependent Ranking of Large Language Models for Peritoneal Dialysis Prescription Review

#### Supplementary Appendix S1. Operational Trap Definitions

**T01, Must: Inadequate Kt/V with declining RKF.** Total weekly Kt/V below 1.7 with declining residual kidney function but unchanged prescription. Guideline basis: ISPD 2020; KDOQI.

**T02, Should: High-transporter long dwell mismatch.** High or high-average transporter exposed to inappropriate long dextrose dwell with poor ultrafiltration or volume status; not coded if icodextrin was appropriately used. Guideline basis: ISPD 2020; Brimble 2006.

**T03, Must: Modified PET ultrafiltration below 400 mL.** 4.25% modified PET ultrafiltration below 400 mL, consistent with ultrafiltration failure or membrane dysfunction. Guideline basis: ISPD 2021.

**T04, Should: Icodextrin not used when indicated.** Long dwell over 8 hours plus high or high-average transporter status with inadequate ultrafiltration or volume overload. Guideline basis: Goossen 2020.

**T05, Should: Modality strategy not revisited.** Persistent or multi-domain PD inadequacy where modality strategy should be reconsidered. Guideline basis: ISPD 2020.

**T06, Optional: High-burden prescription with stable status.** Multiple or high-concentration exchanges despite adequate volume, Kt/V, potassium, and symptoms; potential de-escalation candidate. Guideline basis: ISPD 2020; Maliha 2023.

**T07, Must: Rapid RKF decline with clinical significance.** RKF decline rate at least 0.09 mL/min/month and clinically meaningful adequacy, urine output, volume, hyperkalemia, or preservation implications. Guideline basis: ISPD 2020; CANUSA.

**T08, Must: Persistent hyperkalemia without systematic evaluation.** Recurrent potassium over 5.5 without systematic evaluation of diet, constipation, acidosis, RKF, or PD adequacy. Guideline basis: ISPD 2020; KDOQI.

**T09, Must: Fluid overload with multiple signs.** Edema, elevated blood pressure, weight gain, and multiple antihypertensives indicating volume concern. Guideline basis: ISPD 2020.

**T10, Must: CAPD-to-APD switch with deterioration.** Kt/V or ultrafiltration worsened after transition from CAPD to APD. Guideline basis: ISPD 2020; Brimble 2006.

**T11, Must: Post-peritonitis ultrafiltration decline without repeat PET.** Ultrafiltration declined after peritonitis but PET was not repeated to reassess membrane function. Guideline basis: ISPD 2020.

**T12, Should: Incremental PD not considered.** Substantial residual kidney function preserved but full-dose PD prescribed without documented incremental-PD consideration. Guideline basis: Dong 2019; Qureshi 2023.

**T13, Optional: Biocompatible solution not used when indicated.** Patient eligible for low-GDP solution but standard dextrose continued. Guideline basis: ISPD 2020.

Operational definitions reflect Stage 1 adjudication with Stage 2 prospective confirmation (must-identify kappa 0.92). **Inferential analyses were restricted to must-identify traps** (Stage 1 kappa 0.89; Stage 2 kappa 0.92). Should-consider and optional traps are retained here for descriptive context only and were not used for primary inferential conclusions; optional-tier inter-rater reliability was insufficient for inference (Stage 1 kappa 0.19; Stage 2 kappa 0.14).

**Trap source and severity classification.** All 13 trap categories map to published guidelines or peer-reviewed evidence (ISPD 2020 Practice Recommendations, ISPD 2021 Membrane Dysfunction, KDOQI 2006 commentary, UK Renal Association 2017, Brimble 2006 meta-analysis, Goossen 2020 icodextrin enriched systematic review, Maliha 2023 Kt/V deprescribing, Dong 2019 and Qureshi 2023 incremental PD; the full per-trap source matrix is deposited with the public code release). Most must-identify traps (T01, T03, T07, T08, T09, T10, T11) correspond to long-established hard safety thresholds (Kt/V<1.7, modified PET UF<400 mL, RKF decline rate ≥0.09 mL/min/month, persistent K>5.5 without systematic evaluation, multi-sign fluid overload, post-switch deterioration, post-peritonitis membrane reassessment). Should-consider traps (T02, T04, T05, T12) and optional traps (T06, T13) are guideline-supported but include a stronger expert-judgment component reflecting emerging or context-dependent consensus (e.g., incremental PD eligibility, biocompatible-solution preference, modality-strategy reconsideration), which is consistent with their lower must-identify priority and lower inter-rater reliability.

**Stage 2 per-trap reliability.** Per-trap Cohen's kappa was computed on the 30 Stage 2 cases for every trap on which either rater placed at least one mark. Aggregate Stage 2 trap kappa was 0.77 with must-identify kappa 0.92 (the headline Stage 2 numbers reported in the main manuscript Section 3.2). Traps on which neither rater placed any mark in the Stage 2 sample (T03, T05, T09) are listed as n/a because kappa is undefined when both marginal totals are zero. The full per-case audit trail behind this table is deposited with the project's public code and data release.

| Trap | Tier | Stage 2 $\kappa$ | PI marks | R2 marks |
|---|---|---|---|---|
| T01 | Must | 1.000 | 5 | 5 |
| T02 | Should | 0.714 | 5 | 3 |
| T03 | Must | n/a | 0 | 0 |
| T04 | Should | 0.714 | 5 | 3 |
| T05 | Should | n/a | 0 | 0 |
| T06 | Optional | 1.000 | 1 | 1 |
| T07 | Must | 0.724 | 17 | 19 |
| T08 | Must | 1.000 | 4 | 4 |
| T09 | Must | n/a | 0 | 0 |
| T10 | Must | 0.839 | 4 | 3 |
| T11 | Must | 1.000 | 7 | 7 |
| T12 | Should | 0.464 | 2 | 2 |
| T13 | Optional | 0.000 | 10 | 0 |

*Supplementary Table S1. Stage 2 per-trap Cohen's kappa, n=30 newly generated cases (seed=2027; non-overlapping with Stage 1; PI built gold standards using Stage 1 operational definitions, R2 independently rated blinded to the gold standards). "n/a" indicates that neither rater placed any mark for that trap in the Stage 2 sample, leaving kappa mathematically undefined. T13 kappa of 0.000 reflects persistent R2 under-coding of the biocompatible-solution criterion across both Stage 1 and Stage 2.*

**Per-case agreement.** Of 30 Stage 2 cases, 14 (46.7%) had full agreement on the marked trap set. The persistent R2 under-coding of T13 (biocompatible solution; PI marked 10 cases,

R2 marked 0) was the dominant disagreement pattern across both Stage 1 and Stage 2; the remaining disagreements involved a small number of should-consider trap calls (T02, T04, T12) and overall-judgment-tier mismatches.

### Supplementary Table S2. Model Identity and API Parameters

| Display name | Request model ID | Provider | Context |
|---|---|---|---|
| Claude Sonnet 4.5 | `anthropic/claude-sonnet-4.5` | Anthropic | 200K |
| GPT-5.4 | `openai/gpt-5.4` | OpenAI | 200K |
| Gemini 3.1 Pro | `google/gemini-3.1-pro` | Google | 1M |
| Grok 4.1 Fast | `x-ai/grok-4.1-fast` | xAI | 131K |
| DeepSeek-R1 | `deepseek/deepseek-r1` | DeepSeek | 164K |

**Snapshot identity.** Returned snapshot IDs were captured from the OpenRouter response field `model_id_returned` for all 125 cases in run 1; all 125 (case, run-1) pairs returned identical snapshot strings per model. Returned model IDs were therefore stable within the audited run 1 records; raw provider headers were not separately audited for all 1,875 calls, so this evidence supports model-ID stability rather than a stronger claim about endpoint-level routing consistency. Snapshot IDs: Claude Sonnet 4.5: `anthropic/claude-4.5-sonnet-20250929`; GPT-5.4: `openai/gpt-5.4-20260305`; Gemini 3.1 Pro: `google/gemini-3.1-pro-preview-20260219`; Grok 4.1 Fast: `x-ai/grok-4.1-fast` (no date suffix returned); DeepSeek-R1: `deepseek/deepseek-r1` (no date suffix returned).

**Common parameters.** Temperature 0; maximum output tokens 8192; no streaming; a fixed system-prompt template (registry version v1.0, archived verbatim in Appendix S4 and in the project repository for byte-level verification). All 1,875 calls completed between 2026-05-04 and 2026-05-05 (UTC+8). Each (case, model) pair was queried three times.

**Provider routing and fallback.** OpenRouter requests did not specify provider preference: no `provider.allow_fallbacks`, `provider.order`, or `provider.ignore` options were set in the request body. For Anthropic, OpenAI, and Google models, returned IDs included the provider's date-suffixed snapshot (consistent with routing to the provider's primary endpoint); for Grok and DeepSeek, OpenRouter returned the model ID without a date suffix, indicating that the provider did not surface a versioned snapshot at call time. Two transient request failures during the 1,875-call run were retried successfully against the same model ID; no evidence of model-ID fallback to a different provider was observed in audited records, although raw provider headers were not separately audited for all 1,875 calls.

#### Supplementary Table S3. Additional Reproducibility Notes

Even at temperature 0, deterministic serving stacks can produce output variability (Yuan et al. 2025); triplicate runs were therefore used and run-to-run disagreement is reported in the main manuscript Results section. All raw API responses — including request payload, response body, returned model ID, token usage, elapsed time, and finish reason — are archived per (model, case, run) tuple for byte-level reproducibility audits as part of the public code release described in the Data Availability statement.

### Supplementary Appendix S4. Full LLM System Prompt (verbatim)

The following system-role prompt was sent byte-identically to all five LLMs across all 1,875 calls (registry version v1.0). The case payload was rendered into the user message by a templating routine that strips all gold-standard fields and internal annotations so that no scoring information leaks to the LLM.

```
You are an experienced nephrologist specializing in peritoneal dialysis (PD).
You are reviewing a PD patient's clinical data to evaluate whether their
current PD prescription is appropriate.
Your task:
1. Review all provided clinical data, including PET results, Kt/V, residual
kidney function, ultrafiltration, current PD prescription, longitudinal
trends, medications, and laboratory values.
2. Assess whether the current PD prescription (modality, exchange frequency,
dwell time, dialysate selection) is appropriate for this patient.
3. If you identify any issues, provide specific adjustment recommendations
with clinical rationale.
4. Structure your response as follows:
## Assessment
- Summarize the key clinical findings relevant to prescription adequacy.
## Identified Issues
- List each issue found (if any), with:
  - Description of the problem
  - Supporting data from the case
  - Guideline or evidence basis
## Recommendations
- For each issue, provide specific prescription adjustment recommendations.
- Prioritize recommendations by clinical urgency.
## Reasoning
- Explain your clinical reasoning chain, linking data interpretation to your
conclusions.
```

```
If the current prescription is appropriate, state so explicitly and explain
why.
```

The role framing ("experienced nephrologist") is consistent with the parent CKD-LLM series. Structured output sections enable automated extraction and downstream scoring against the gold standard; the extraction and scoring code is part of the public code release.

### Supplementary Appendix S5. Weighted Clinical Score (WCS) Rubric

The Weighted Clinical Score is a deterministic per-output composite over four clinical dimensions, computed by the scoring code against per-case rubrics stored as machine-readable gold-standard files (one JSON per case). The full JSON schema is provided in the public code release.

**Formula.**

$$\mathrm{WCS} = \sum_{d \in \{\mathrm{Saf},\mathrm{Diag},\mathrm{Tx},\mathrm{Reason}\}} w_d \cdot \frac{\max(0,\ \min(\mathrm{raw}_d,\ \mathrm{cap}_d))}{\mathrm{cap}_d}$$

where $w = (0.35,\ 0.25,\ 0.25,\ 0.15)$ and $\mathrm{cap}_d = 5$ points per dimension. WCS is bounded in $[0, 1]$. The dimension weights were chosen *a priori* to reflect the relative clinical importance of safety over reasoning style; see Section S6 for sensitivity analysis under alternative weightings.

**Per-dimension scoring.** Each dimension's raw score is the sum of awarded criterion points, where every criterion has a polarity (`positive`: award if concept present in LLM output; `negative`: deduct if concept present, i.e., the LLM did the unsafe thing). The dimension is then clipped at $[0, 5]$.

**Safety (weight 0.35, max 5).** Positive criteria (representative): correctly recognizes hyperkalemia management hierarchy; flags fluid overload signs; recommends repeat PET after peritonitis; identifies UF failure thresholds. Negative criteria: recommends an unsafe dose escalation when RKF is collapsing; suggests stopping PD without bridging plan; misses critical electrolyte abnormality.

**Diagnostic Accuracy (weight 0.25, max 5).** Positive criteria: correctly interprets transporter category from D/P creatinine; identifies inadequate Kt/V relative to target; recognizes incremental-PD eligibility; correctly attributes UF decline to membrane dysfunction. Negative criteria: misclassifies transporter status; quotes wrong adequacy threshold; confuses APD and CAPD parameters.

**Treatment Appropriateness (weight 0.25, max 5).** Positive criteria: prescription change matches the trap (e.g., adds icodextrin for high-transporter long-dwell mismatch; switches CAPD→APD when indicated); cites guideline-supported dosing; integrates RKF preservation strategy. Negative criteria: recommends an off-guideline intervention; suggests unnecessary modality switch.

**Reasoning Quality (weight 0.15, max 5).** Positive criteria: links data points to conclusions explicitly; references appropriate guideline; uses tier-appropriate uncertainty language. Negative criteria: confabulates a guideline citation; reasons from absent or contradictory data.

**Concept matching.** Each criterion is evaluated by a normalized concept match (case-insensitive substring, whitespace-collapsed) against the LLM output. Criterion lists are case-specific: every gold-standard JSON encodes the criteria for that case (median 4–6 criteria per dimension). The concept-match function is shared with the trap-detection scorer to ensure internal consistency.

**Limitations of the rubric.** (i) The rubric rewards literal lexical presence of expected concepts, which can disadvantage outputs that paraphrase correctly using synonyms not anticipated in the criterion list. (ii) Negative-polarity criteria are rare in practice and cap dimension scores at zero; severe unsafe outputs are flagged separately as judgment-level inappropriateness. (iii) The four-dimension structure with these specific weights was a pre-specified design choice; weight robustness is tested in Appendix S6.

### Supplementary Box S1. Mechanistic Case Audits: How WCS Rubric Behavior Produced Observed Scores

These three case audits are presented as **mechanistic explanations of WCS rubric behavior**, not as a representative sample documenting the magnitude of WCS–expert disagreement (which would require a larger blinded panel; see main manuscript Limitations). Each audit traces how literal concept matching produced its observed score for that specific output, given the embedded trap set, the LLM response structure, and the expert raters' clinical reasoning. The three audits jointly illustrate the two scoring directions identified in Discussion §4.1: (i) compact paraphrase scoring low on lexical match but high on expert assessment, and (ii) extensive enumeration scoring high on lexical match but only moderately on expert assessment. Each case was rated independently by two board-certified nephrologists blinded to model identity, gold standard, and WCS.

**Vignette 1: Grok on PD-097 (WCS 0.05; expert mean 92).** PD-097 embeds T07 (rapid RKF decline; residual GFR 2.9 mL/min, residual Kt/V 0.36, urine output 351 mL/day at 19 PD-months) and T13 (biocompatible-solution eligibility for a 55-year-old polycystic-kidney patient on standard dextrose). Grok produced a concise output that correctly named the RKF-decline pattern, recommended escalation of total Kt/V toward the 1.7 target, and explicitly flagged the need for repeat ultrafiltration assessment, but used short paraphrases (e.g., "urine clearance is dropping", "review long-term solution choice") that did not contain the literal criterion strings encoded in the GS rubric. Both nephrologists rated the output 90 and 95 because it captured the principal safety concern compactly. The WCS rubric counted very few literal matches across all four dimensions, producing 0.05.

**Vignette 2: Claude on PD-021 (WCS 0.23; expert mean 92.5).** PD-021 embeds T01 (inadequate Kt/V given declining RKF) and T02 (high-transporter long-dwell mismatch with poor ultrafiltration). Claude generated a long output (>20 issues raised) that nested the two embedded traps inside an enumerated list of 15+ secondary concerns (e.g., bone-mineral metabolism, peritonitis prophylaxis, lipid management). Both nephrologists rated the output 90 and 95 because the principal traps were correctly identified with appropriate management; however, the dilution of relevant content across many issues caused the WCS criterion-matching pass to score weakly on diagnostic-accuracy and treatment-appropriateness dimensions, yielding 0.23.

**Vignette 3: Claude on PD-049 (WCS 0.89; expert mean 79).** PD-049 embeds T05 (modality strategy not revisited despite persistent inadequacy) and T13 (biocompatible solution). Claude produced a long, well-structured output that hit nearly every literal criterion in the four-dimension WCS rubric, scoring 0.89. Both nephrologists rated the output 78 and 80 because the recommendations, while complete in coverage, did not prioritize the principal modality-strategy decision and therefore lacked the clinical focus they expected; they noted "thorough but does not lead with the key decision". The WCS scored coverage; expert ratings scored prioritization.

**Interpretation.** Across these three cases, WCS rewarded literal multi-criterion lexical coverage and penalized concise paraphrase. Expert ratings instead weighed whether the principal trap was correctly identified and whether the output prioritized that finding. The dissociation at $\rho \approx -0.17$ is consistent with this difference.

### Supplementary Appendix S6. WCS Weight Sensitivity Analysis

To test whether the primary WCS weighting (Safety 0.35 / Diagnostic 0.25 / Treatment 0.25 / Reasoning 0.15) drives the model-ranking conclusion, we re-computed per-case WCS under three alternative pre-specified schemes and compared model rankings and expert-correlation results across all four. Analysis code and result files are part of the public code release; n = 1,875 (case × model × run) records.

**Schemes tested.**

| Scheme | Safety | Diagnostic | Treatment | Reasoning |
|---|---|---|---|---|
| Primary | 0.35 | 0.25 | 0.25 | 0.15 |
| Equal | 0.25 | 0.25 | 0.25 | 0.25 |
| Safety-heavy | 0.50 | 0.20 | 0.20 | 0.10 |
| Reasoning-up | 0.30 | 0.20 | 0.20 | 0.30 |

**Mean WCS per model under each scheme.**

| Model | Primary | Equal | Safety-heavy | Reasoning-up |
|---|---|---|---|---|
| Claude Sonnet 4.5 | 0.665 | 0.659 | 0.664 | 0.650 |
| GPT-5.4 | 0.581 | 0.565 | 0.583 | 0.550 |
| Gemini 3.1 Pro | 0.498 | 0.486 | 0.494 | 0.471 |
| DeepSeek-R1 | 0.486 | 0.458 | 0.495 | 0.437 |
| Grok 4.1 Fast | 0.448 | 0.434 | 0.443 | 0.415 |

**Rank order (1 = highest WCS) across schemes.**

| Model | Primary | Equal | Safety-heavy | Reasoning-up |
|---|---|---|---|---|
| Claude Sonnet 4.5 | 1 | 1 | 1 | 1 |
| GPT-5.4 | 2 | 2 | 2 | 2 |
| Gemini 3.1 Pro | 3 | 3 | 4 | 3 |
| DeepSeek-R1 | 4 | 4 | 3 | 4 |
| Grok 4.1 Fast | 5 | 5 | 5 | 5 |

**Kendall's coefficient of concordance.** Across the four schemes, W = 0.963 (chi-square 15.40, df 4, p = 0.004), indicating that the WCS-derived ranking is extremely stable across weight choice. The only rank change observed was a single adjacent swap between DeepSeek-R1 and Gemini 3.1 Pro under the safety-heavy scheme; the top two ranks (Claude, GPT-5.4) and the bottom rank (Grok) were identical under every scheme.

**Expert correlation under each scheme** (n = 25 blinded sub-study; Spearman $\rho$ with percentile bootstrap 95% CI, $n_{boot}$ = 10,000, seed 42).

| Scheme | n | Spearman $\rho$ | 95% CI | p |
|---|---|---|---|---|
| Primary | 25 | −0.173 | [−0.586, 0.287] | 0.41 |
| Equal | 25 | −0.141 | [−0.537, 0.293] | 0.50 |
| Safety-heavy | 25 | −0.143 | [−0.573, 0.325] | 0.50 |
| Reasoning-up | 25 | −0.083 | [−0.488, 0.350] | 0.69 |

**Interpretation.** Model rankings were stable across the four weight schemes (Kendall's W = 0.963; only one adjacent swap), and the WCS–expert correlation was small and non-significant under every scheme. The dissociation reported in the main manuscript is therefore not an artifact of the primary 0.35/0.25/0.25/0.15 weighting.

### Supplementary Appendix S7. WCS–Expert Subset Sensitivity Analysis

To test whether the WCS-versus-expert null correlation in the n=25 sub-study depends on case complexity, the choice of WCS dimension, the upper-tail compression of expert ratings, or a single high-verbosity model, we recomputed Spearman $\rho$ under four pre-specified subset restrictions. Analysis code and result files are part of the public code release.

| Subset | Definition | n | Spearman $\rho$ | 95% CI | p |
|---|---|---|---|---|---|
| Full sample (reference) | all rated outputs | 25 | $-0.173$ | $[-0.586, 0.287]$ | 0.41 |
| (a) Must-identify-trap cases only | case contains $\geq$1 must trap | 19 | $-0.249$ | $[-0.697, 0.280]$ | 0.30 |
| (b) Safety dimension only | WCS = Safety/5 | 25 | $-0.097$ | $[-0.503, 0.354]$ | 0.65 |
| (c) Expert-rating trimmed 10% | drop top/bottom 10% rating | 19 | $-0.319$ | $[-0.724, 0.203]$ | 0.18 |
| (d) Exclude Claude outputs | remove most-verbose model | 20 | $+0.013$ | $[-0.477, 0.510]$ | 0.96 |

**Interpretation.** None of the four restrictions produced a positive WCS–expert association; all five $\rho$ estimates ranged from $-0.32$ to $+0.01$ with 95% CIs spanning zero. Excluding Claude moved $\rho$ from $-0.17$ to $+0.01$, consistent with Claude's literal-coverage WCS scores over-stating perceived clinical adequacy compared with expert ratings. These exploratory analyses do not exclude all positive association; a larger expert panel is required for confirmatory inference.
